# Prognostic nutrition index, neutrophil-to-lymphocyte ratio and platelet-to-lymphocyte ratio combined detection, superior to single detection correlates with prognosis of patients with chronic hepatitis C-associated cirrhosis complicated by T2DM

**DOI:** 10.1101/2022.12.30.22284073

**Authors:** Dong Wu, Xiaowu Wang, Yunyun Ding, Yan Liu, Tuantuan Li, Yi lang Zhu, Yong Gao, Xiaojuan Wang

**Author notes:** **Correspondence** Xiaojuan Wang, Department of Pharmacy, Fuyang People’s Hospital, Fuyang, Anhui, China. These authors contributed equally to this work (Dong Wu, Xiaowu Wang). **Funding:** The author(s) received no specific funding for this work. **Competing interests:** The authors have declared that no competing interests exist.

## Abstract

**Background:** Prognostic nutrition index (PNI), Neutrophil-to-Lymphocyte Ratio (NLR) and Platelet-to-Lymphocyte Ratio (PLR) have been studied widely in cancer diseases. However, their correlation with chronic hepatitis C-associated cirrhosis complicated by T2DM mellitus is unknown.

**Aim:** To investigate the correlation of PNI, NLR and PLR with chronic hepatitis C-associated cirrhosis complicated by T2DM.They were associated with the prognosis of patients.

**Methods:** We investigated 226 patients. Of the patients, 56 cases were with chronic hepatitis C-associated cirrhosis complicated by T2DM mellitus patients (group A), 85 cases were with chronic hepatitis C-associated cirrhosis patients (group B), and 85 cases were with T2DM patients (group C). According to the prognosis of chronic hepatitis C-associated cirrhosis complicated by T2DM after 6 months of treatment, patients were divided into poor prognosis (23 cases) and good prognosis (33 cases). The baseline data and of all patients were analyzed. All data were collected from the database of the hospital patient electronic medical record system.

**Results:** A comparison of baseline data among the three groups showed significant differences in age (*P* value=0.008). The levels of PNI were obvious difference in three groups (*P* value < 0.01). The level of NLR in the group A was significantly lower than in the group C. The level of NLR in the group B was significantly lower than in the group C and the group A. The level of PNI in the group A was significantly lower than in the group C. The level of PNI in the group A was significantly lower than in the group B. The level of PLR in the group A was significantly lower than in the group C. The level of PLR in the group B was significantly higher than in the group C. NLR, PNI, PLR were significantly different in the good prognosis group and the poor prognosis group (*P* value < 0.05). We calculated AUC for combined determination of PNI, NLR and PLR, and it showed excellent diagnostic performance (AUC = 0.911,95% CI 0.741-0.985, Sensitivity = 80.00 %, and Specificity = 88.89%).

**Conclusions:** PNI, NLR and PLR were closely related to the prognosis of chronic hepatitis C-associated cirrhosis complicated by T2DM, and their combined detection had the highest specificity and sensitivity for early prediction of the poor prognosis of chronic hepatitis C-associated cirrhosis complicated by T2DM, which had important clinical value.

## 1 Introduction

Chronic hepatitis C (CHC) is an inflammatory liver disease caused by the hepatitis C virus (HCV), which is a major public health problem and a leading cause of chronic liver disease. With its characteristic high degree of chronicity, HCV infection often causes chronic inflammatory necrosis of the liver, which leads to liver cirrhosis and even hepatocellular carcinoma (HCC) ^[1]^. Although acute HCV infection is usually not life-threatening, all complications such as ascites, upper gastrointestinal bleeding secondary to varix, and portal hypertensive gastropathy, which are associated with chronic hepatitis, will indicate the decreased quality of life and a poor prognosis ^[2]^. At the same time, extrahepatic manifestations will occur gradually, which include autoimmune disorders, mixed cryoglobulinemia, Sjogren’s syndrome, and endocrinological diseases such as autoimmune thyroid disorders and type 2 diabetes (T2DM) ^[3]^. HCV infection has been shown to be a significant risk factor for developing T2DM. CHC, cirrhosis, and decompensated cirrhosis contribute to an increasingly greater risk of T2DM in individuals with HCV, but HCV clearance spontaneously or through clinical treatment may immediately reduce the risk of the onset and development of T2DM ^[4]^. At present, it is clinically infected with HCV virus, and the prevalence of T2DM is generally higher than that of the normal population. Also, studies in liver cancer patients on the platelet-to-lymphocyte ratio (PLR) and neutrophil-to-lymphocyte ratio (NLR), which are two important indicators of systemic inflammation and have been shown to be prognostic parameters in various cancer treatments ^[5-7]^. The PNI, which was calculated based on the serum albumin and circulating peripheral blood lymphocyte count, has been used to assess the immune nutritional status of cancer patients. Prognostic nutrition index (PNI) had also been verified as a useful prognostic biomarker in various cancers, including esophageal carcinoma and osteosarcoma ^[8-9]^. In CHC patients, the albumin is also regarded as the important factor for the liver function.

Although increasing evidence shows that the PNI, NLR and PLR, which can inflammation and nutritional statuses, can accurately predict cancer patient prognosis, the relationship between PNI, NLR and PLR, and clinical Prognosis in patients with chronic hepatitis C-associated cirrhosis complicated by T2DM remains unclear and has not been verified. The main purpose of this study was to evaluate the prognostic value of PNI, NLR and PLR in chronic hepatitis C-associated cirrhosis complicated by T2DM patients.

## 2 Materials and methods

### 2.1 Study subjects

Following the application of inclusion and exclusion criteria, 56 chronic hepatitis C-associated cirrhosis complicated by T2DM patients (group A), 85 chronic hepatitis C-associated cirrhosis patients (group B), and 85 T2DM patients (group C) from the Fu yang Second People’s Hospital were included in the study from January, 2018 to December, 2021. Hepatitis C-associated cirrhosis were diagnosed according to clinic standards by laboratory parameters and liver histopathology tests. According to the prognosis of chronic hepatitis C-associated cirrhosis complicated by T2DM after 6 months of treatment, patients were divided into poor prognosis group (23 cases) and good prognosis group (33 cases). Permission to conduct the study was approved by the Ethics Committee of Fu yang Second People’s Hospital; and informed written consent was obtained from each patient.

### 2.2 Clinical laboratory data

Blood biochemistry parameters serum albumin (ALB), aspartate aminotransferase (AST) and alanine aminotransferase (ALT) were measured using HITACHI 7600-020 automated biochemistry analyzer. The platelet count, lymphocyte count, mononuclear count, and neutrophil counts were analyzed by an XE-2100 hematology analyzer (Sysmex, Kobe, Japan). The NLR, PNI and PLR were calculated as follows: NLR = neutrophil count/lymphocyte count; PNI= ALB(g/L) + 5×total lymphocyte count (10^9^/L); PLR=platelet count/lymphocyte count. All patients used data from their first laboratory test on admissions. All the operations were done by specially□assigned personnel and in strict accordance with the instructions regarding the use of the reagents.

### 2.3 Statistical Analysis

The statistical analysis was performed using IBM SPSS Statistics version 22.0 software. Summary statistics of the study population were expressed as the mean ± standard deviation (mean±SD) or median value with the interquartile range (IQR), as appropriate. The Kolmogorov-Smirnov test was performed to evaluate variable distribution. Comparisons of demographic and clinical parameters of the two groups were performed using the Chi-square test, Student t-test (independent samples t-test), or Mann-Whitney U-test, as appropriate; the Kruskal-Wallis-test was used for the comparison of more than two groups. A receiver operating characteristic (ROC) curve was used for the evaluation of predictors and to determine their sensitivities and specificities. A two□sided P value less than 0.05 was considered significant. The results of the analysis were obtained using SPSS for windows.

## 3 Results

### 3.1 Baseline data

The study involved 226 patients. Sex, age, region, alcohol consumption characteristics of patients were summarized in **Table 1**. Of the patients, 56 cases were with chronic hepatitis C-associated cirrhosis complicated by T2DM patients (group A), 85 cases were with chronic hepatitis C-associated cirrhosis patients (group B), and 85 cases were with T2DM patients (group C). There were not significant differences in sex among the three groups(*P* value>005). There were significant differences in age, region, alcohol consumption among the three groups (*P* value<0.05).

**Table 1.**
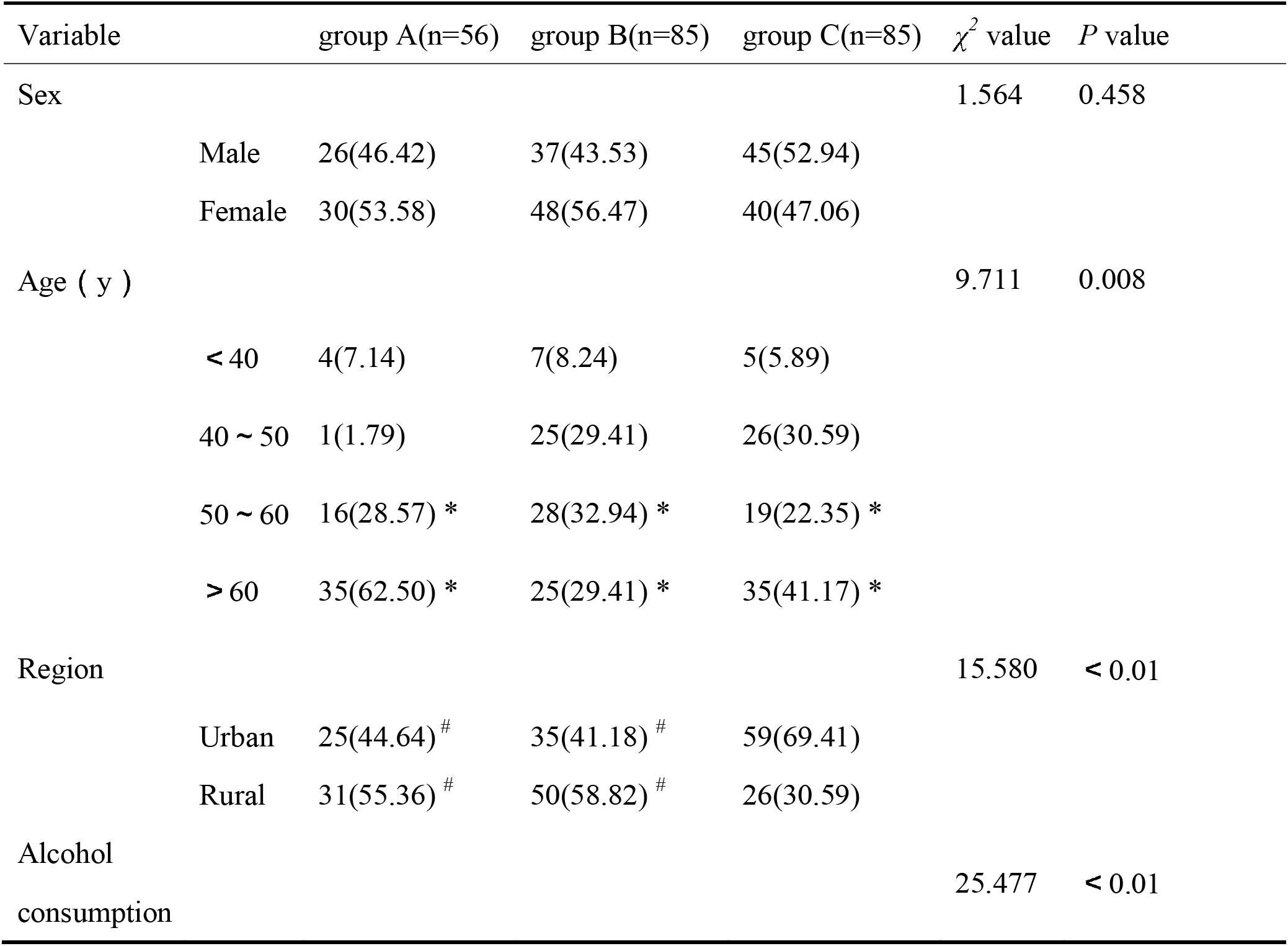
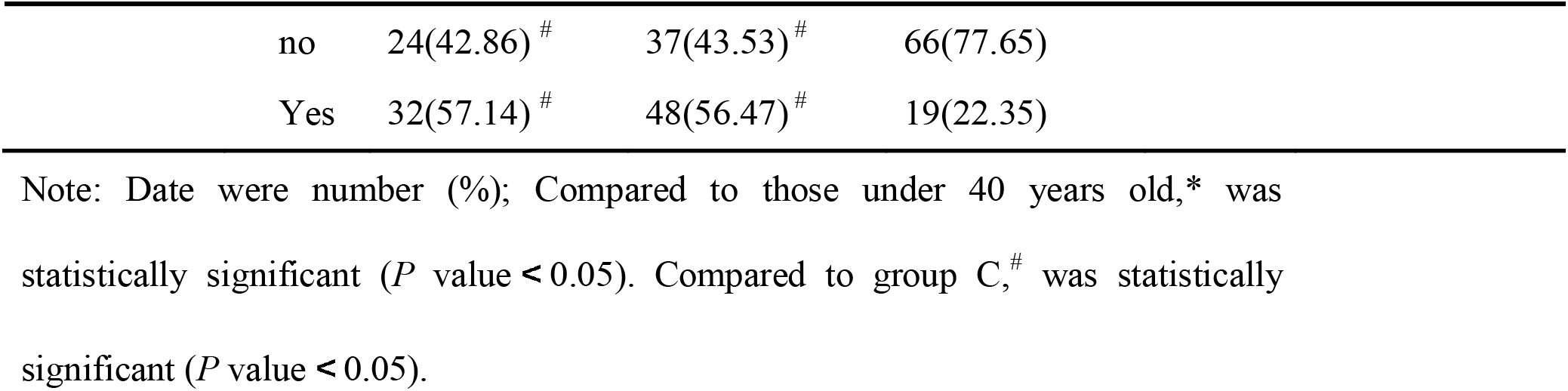
Baseline characteristics of patients with chronic hepatitis C-associated cirrhosis complicated by T2DM

### 3.2 Clinical laboratory data

The Clinical laboratory data of the patients were shown in **Table 2**. The levels of ALT,AST, NLR, PLR were significantly different in three groups (*P* value < 0.05).The levels of PNI were obvious difference in three groups (*P* value < 0.01).The level of ALT and AST in the group A [median (P25, P75): 46.00(21.00,68.00) U/L; 28.00(26.00,55.00) U/L; 0.17(0.14,0.26)] was significantly higher than in the group C [median (P25, P75): 15.00(14.00,30.00) U/L; 11.00(15.00,30.00) U/L; 0.12(0.08,0.15)]. The level of ALT and AST in group B [median (P25, P75): 49.00(19.00,69.00) U/L; 67.00(27.00,93.00) U/L; 0.19(0.15,0.24)] was significantly higher than in the group C. But The level of ALT and AST in the group A was not significantly compared with in the group B. The level of NLR in the group A (mean ± SD: 1.94±1.42) was significantly lower than in the group C (mean ± SD: 3.51±4.78). The level of NLR in the group B (mean ± SD: 1.24±0.40) was significantly lower than in the group C and the group A. The level of PNI in the group A [median (P25, P75): 46.35(42.66,54.54)] was significantly lower than in the group C [median (P25, P75): 50.65(47.20,53.73)]. The level of PNI in the group A was significantly lower than in the group B [median (P25, P75): 48.85(43.00,50.99)]. But The level of PNI in the group B was not significantly compared with in the group C. The level of PLR in the group A [median (P25, P75): 111.45±80.05] was significantly lower than in the group C [median (P25, P75): 147.51±101.50]. The level of PLR in the group B [median (P25, P75): 91.86±41.62] was significantly higher than in the group C. But The level of PLR in the group A was not significantly compared with in the group B.

**Table 2.**
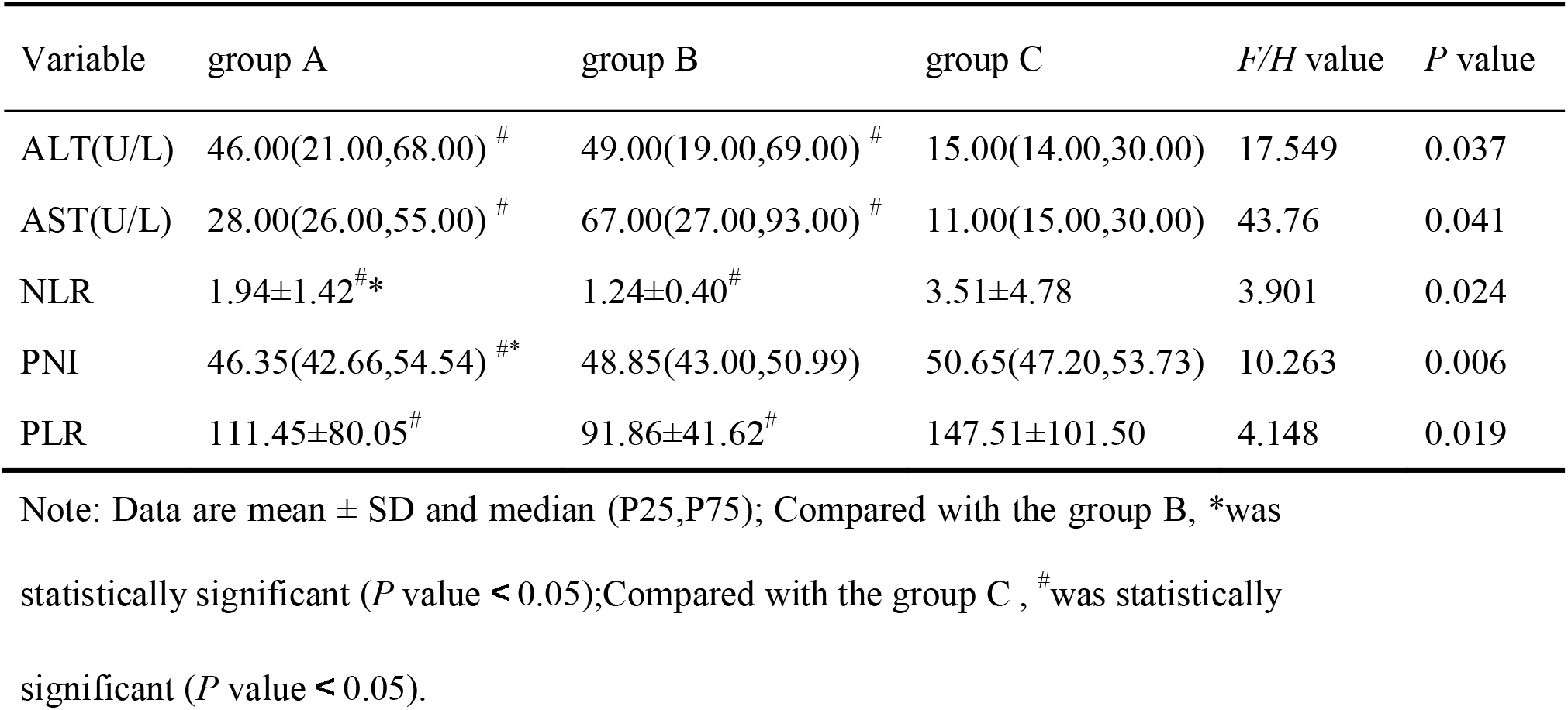
Clinical laboratory data between groups

### 3.3 Prognostic significance

Prognostic significance of laboratory indicators of chronic hepatitis C-associated cirrhosis complicated by T2DM patients (group A) were shown in **Table 3**. Group A was divided into good prognosis group (n=23) and poor prognosis group(n=33). NLR, PNI, PLR were significantly different in the good prognosis group and the poor prognosis group (*P* value<0.05). The level of NLR in the poor prognosis group (mean ± SD: 2.49±0.71) was higher significantly than in the good prognosis group (mean ± SD: 1.14±1.99) (t=1.914, *P* value=0.022). The level of PNI in the poor prognosis group [median (P25, P75): 42.65(37.66,49.54)]was lower significantly than in the good prognosis group [median (P25, P75): 49.35(42.66,54.54)] (z=-4.322, *P* value < 0.001). The level of PLR in the poor prognosis group (mean ± SD: 94.57±35.27) was lower significantly than in the good prognosis group (mean ± SD: 142.45±69.22) (t=2.409, *P* value=0.023). But The level of ALT and AST in the good prognosis group was not significantly compared with in the poor prognosis group (z=-1.128, *P* value=0.259; z=-1.043, *P* value=0.307; z=-0.729, *P* value=0.472).

**Table 3.**
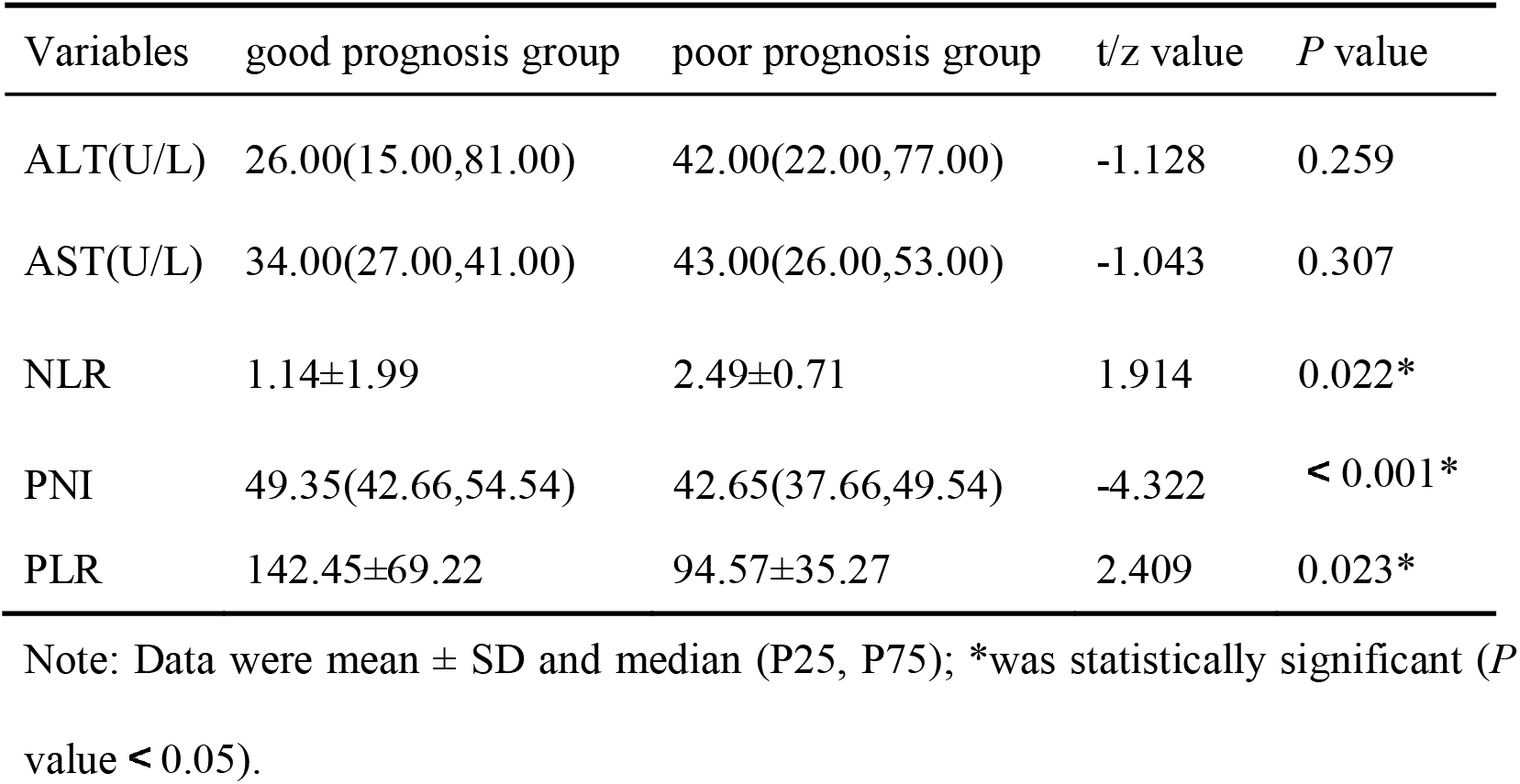
Prognostic significance of laboratory indicators

### 3.4 ROC curve analysis

The ROC curve was used to analyze the prognosis of chronic hepatitis C-associated cirrhosis complicated by T2DM patients. ROC curve analysis illustrated that the AUCs of PNI, NLR, PLR for Evaluation of prognosis of chronic hepatitis C-associated cirrhosis complicated by T2DM mellitus patients were 0.872 (95% CI 0.692 to 0.967), 0.739 (95% CI 0.539 to 0.885), 0.717 (0.516 to 0.869) respectively. When PNI and NLR were combined, the AUC was 0.906 (95% CI 0.734 to 0.983). When PNI and PLR were combined, the AUC was 0.906 (95% CI 0.734 to 0.983). When NLR and PLR were combined, the AUC was 0.739 (95% CI 0.539 to 0.885). We calculated AUC for combined determination of PNI, NLR and PLR, and it showed excellent diagnostic performance (AUC = 0.911,95% CI 0.741-0.985, Sensitivity = 80.00 %, and Specificity = 88.89%) (**Table 4 and Figure 1**).

**Table 4.**
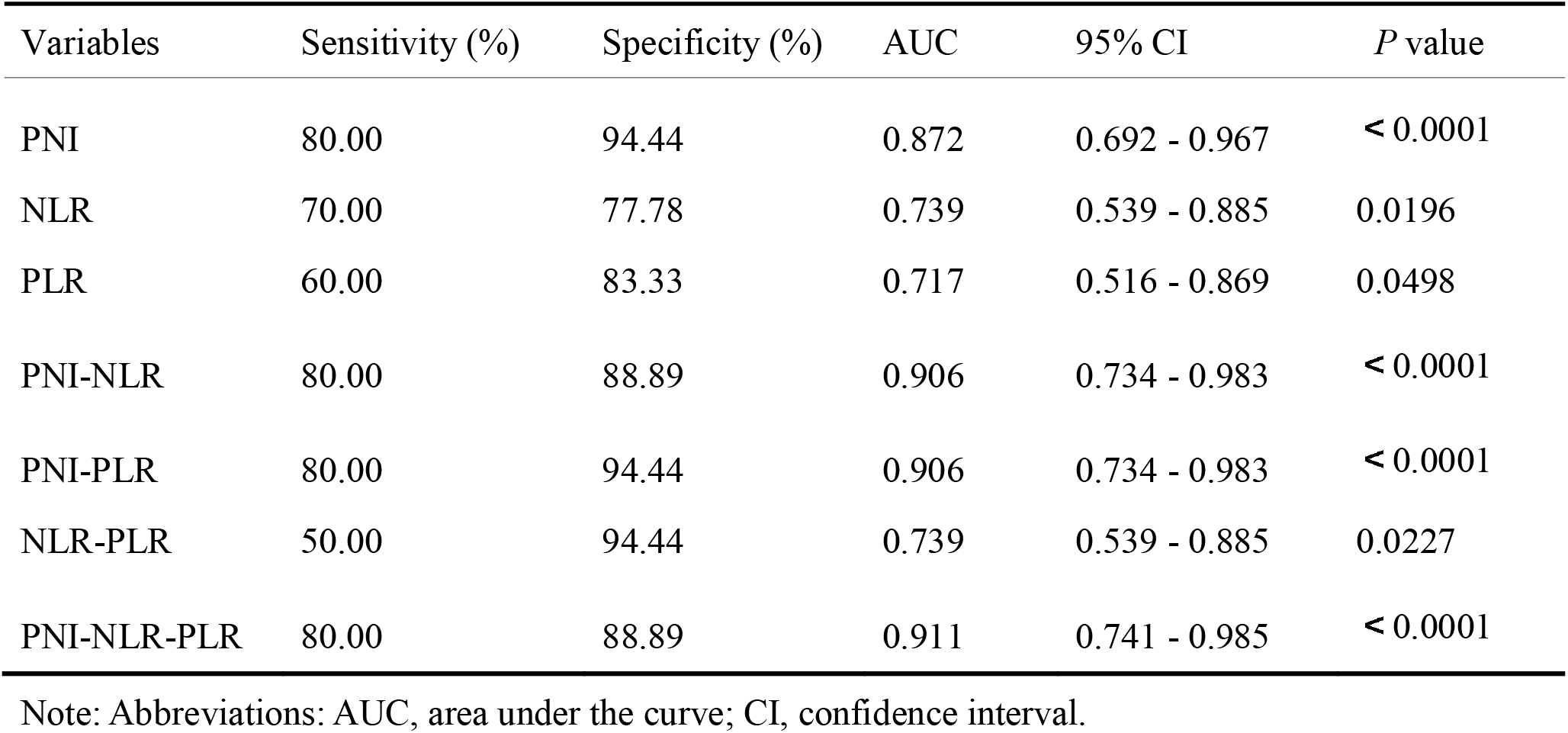
ROC curve analysis of clinical laboratory indicators

| Variables | Sensitivity (%) | Specificity (%) | AUC | 95% CI | *P* value |
| --- | --- | --- | --- | --- | --- |
| PNI | 80.00 | 94.44 | 0.872 | 0.692 - 0.967 | < 0.0001 |
| NLR | 70.00 | 77.78 | 0.739 | 0.539 - 0.885 | 0.0196 |
| PLR | 60.00 | 83.33 | 0.717 | 0.516 - 0.869 | 0.0498 |
| PNI-NLR | 80.00 | 88.89 | 0.906 | 0.734 - 0.983 | < 0.0001 |
| PNI-PLR | 80.00 | 94.44 | 0.906 | 0.734 - 0.983 | < 0.0001 |
| NLR-PLR | 50.00 | 94.44 | 0.739 | 0.539 - 0.885 | 0.0227 |
| PNI-NLR-PLR | 80.00 | 88.89 | 0.911 | 0.741 - 0.985 | < 0.0001 |

Note: Abbreviations: AUC, area under the curve; CI, confidence interval.

**Figure 1.**
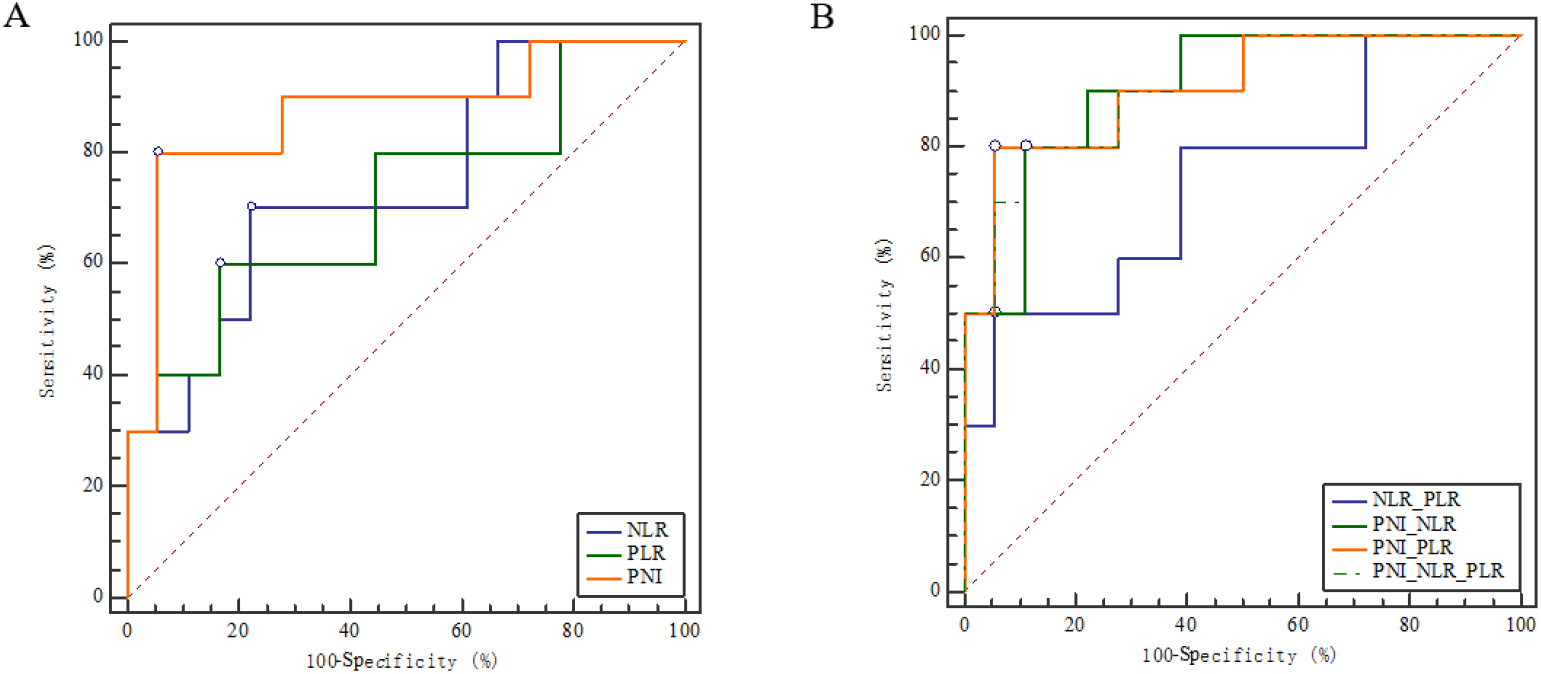
Receiver operator characteristic curves comparing the potential of different variables to predict the prognostic value of combined detection in patients with chronic hepatitis C-associated cirrhosis complicated by T2DM.

## 4 Discussion

HCV is a risk factor for cirrhosis. Cirrhosis takes an average of 30 years to develop after the HCV infection, although the average period varies considerably from person to person. Cirrhosis is the cause of 4% annual deaths worldwide ^[10]^. Chronic hepatitis C-associated cirrhosis complicated by T2DM the pathogenesis is relatively complex, acute and chronic hyperglycemia. A proinflammatory state, it can promote the synthesis and synthesis of various inflammatory factors Secretion, increase liver oxidative stress response, and then participate in liver inflammation Reaction and fibrosis progression.

Liver cirrhosis is the terminal stage of most chronic liver conditions, with a high risk of mortality. Careful evaluation of the prognosis of cirrhotic patients and providing precise management are crucial to reduce the risk of mortality ^[11]^. In recent years, continuous efforts have been made to investigate the prognostic value of body fluid biomarkers for cirrhotic patients, and promising results have been reported. Since the collection of fluid specimens is easy, noninvasive, and repeatable, fluid biomarkers can be ideal indicators to predict the prognosis of cirrhosis ^[11]^. In the present study, we evaluated and compared the performances of PNI, PLR and NLR in chronic hepatitis C-associated cirrhosis complicated by T2DM patients.

Chronic hepatitis C-associated cirrhosis complicated by T2DM patients was higher in rural areas and the proportion of alcohol consumption was also higher compared with T2DM patients without chronic hepatitis C-associated cirrhosis. Similarly, a higher proportion of alcohol consumption were found in patients with chronic hepatitis C-associated cirrhosis without T2DM. Compared with T2DM with or without chronic hepatitis C-associated cirrhosis, the prevalence of chronic hepatitis C-associated cirrhosis patients in rural areas is higher than that in urban areas, and the number of alcohol consumption is more than that in no alcohol consumption. It suggested that attention should be paid to the prevention and treatment of hepatitis C in rural drinking patients. The results were that the number of patients with chronic hepatitis C-associated cirrhosis complicated by T2DM patients in the age group over 60 years old and 50-60 years old were more than that in the age group under 40 years old, and the difference were statistically significant (P < 0.05), indicating that the older the patients with chronic hepatitis C-associated cirrhosis were, the more likely they were to be complicated withT2DM. At the same time, the author found that the number of patients in the age group over 60 years old and the age group between 50 and 60 years old T2DM patients with chronic hepatitis C-associated cirrhosis was also higher than that in the age group under 40 years old, and the difference was statistically significant (P < 0.05). NguyenC T et al. ^[12]^ also confirmed this view through Meta analysis. The increase of age was positively correlated with the prevalence of T2DM, indicating that the incidence of T2DM gradually increases with the increase of age, suggesting that age was a risk factor for T2DM, and the older people need to pay more attention to the management of blood sugar.

Ali S A et al. ^[13]^ showed that Serum levels of liver enzymes (AST, ALT) in both HCV and HCV/DM groups were significantly higher than the control group, but without any discrimination among the two HCV diseased groups. HCV infection would be in the human liver cells and replicated in large numbers, resulting in the release of enzymes in the cytoplasm into the bloodstream, so that the activity of these enzymes in the serum increased, causing liver damage, liver function abnormalities in patients ^[14]^. Our results showed that the values of ALT and AST in chronic hepatitis C-associated cirrhosis patients were higher than those in T2DM patients without chronic hepatitis C-associated cirrhosis, and the difference was statistically significant (P<0.05). Therefore, the liver function of chronic hepatitis C-associated cirrhosis patients was worse than that of T2DM patients without chronic hepatitis C-associated cirrhosis, and attention should be paid to the protection of liver in chronic hepatitis C-associated cirrhosis patients and prevent further deterioration and induce a series of extrahepatic manifestations.

Proposed inflammatory scores, such as NLR and PLR have been considered as useful indicators for predicting the prognosis and survival in various cancers. However, due to variance in study designs and sample sizes, these studies have reported inconsistent results ^[15]^. Our results showed that the values of NLR and PLR in chronic hepatitis C-associated cirrhosis patients were lower than those in T2DM patients without chronic hepatitis C-associated cirrhosis, and the difference was statistically significant (P<0.05). NLR and PLR were significantly different in the good prognosis group and the poor prognosis group (*P* value<0.05). A previous study by Li et al. showed PLR was elevated in cirrhosis patients compared to that in liver disease, being consistent with our study ^[16]^. From the pathogenetic analysis, hypersplenism and bone marrow suppression caused by hepatitis virus infection in patients with cirrhosis lead to decreased platelet count and decreased lymphocytes. In this study, compared with chronic hepatitis C-associated cirrhosis without T2DM patients, the decline of platelets in chronic hepatitis C-associated cirrhosis withT2DM patients was more significant than that of lymphocytes. For the prognosis of patients with hepatitis C cirrhosis complicated with diabetes, the degree of platelet decline in patients with poor prognosis was more significant than that of lymphocytes. NLR indicates the balance of the inflammatory and immune systems, making the NLR a useful index that reflects systemic inflammation responses ^[17]^. The NLR has attractive prognostic value for patients with hepatopathy. Our results showed that the values of NLR in the good prognosis group were lower than those in the poor prognosis group, and the difference was statistically significant (P< 0.05). ROC curve analysis illustrated that the AUCs of NLR and PLR for Evaluation of prognosis of chronic hepatitis C-associated cirrhosis complicated by T2DM mellitus patients were 0.739 (95% CI 0.539 to 0.885), 0.717 (0.516 to 0.869) respectively. But the AUCs of NLR and PLR were below 0.750, thus leading to poor predictive value.

The concept of the PNI was first proposed by Buzby et al. ^[18]^ to evaluate the risk of gastrointestinal surgery. Later, it was also used in prognostic studies of various diseases. The relationship between PNI and clinical Prognosis in patients with chronic hepatitis C-associated cirrhosis complicated by T2DM remains unclear and has not been verified. Our results showed that the level of PNI in the chronic hepatitis C-associated cirrhosis complicated by T2DM patients was significantly lower than in the chronic hepatitis C-associated cirrhosis patients. The level of PNI in the chronic hepatitis C-associated cirrhosis without T2DM patients was significantly lower than in the T2DM patients. But The level of PNI in the chronic hepatitis C-associated cirrhosis patients with T2DM was not significantly compared with in the chronic hepatitis C-associated cirrhosis patients without T2DM. The level of PNI in the poor prognosis group was lower significantly than in the good prognosis group.The liver was the main place for the metabolism of sugars, when sugars metabolize work When it was damaged, other functions of the liver were often affected. The PNI, which was calculated based on the serum albumin and circulating peripheral blood lymphocyte count, has been used to assess the immune nutritional status of cancer patients. Many T2DM patients had an overactive immune system, meaning they had chronic inflammation in their bodies. The inflammatory state could be led to impaired liver function. PNI can evaluate the nutritional status and prognosis of patients. ROC curve analysis illustrated that the AUC of PNI for Evaluation of prognosis of chronic hepatitis C-associated cirrhosis complicated by T2DM mellitus patients were 0.872 (95% CI 0.692 to 0.967). the AUC of PNI were more than 0.750, thus leading to good predictive value. But AUC for combined determination of PNI, NLR and PLR, and it showed excellent diagnostic performance (AUC = 0.911,95% CI 0.741-0.985, Sensitivity = 80.00 %, and Specificity = 88.89%) which also confirmed the high prediction efficiency.

This study has several limitations. The sample size was relatively small. Since this study was a retrospective study, not all patients were continuously monitored for all indicators in the blood including PNI.

## Conclusions

Findings suggest that PNI, NLR and PLR levels can be used to estimate the prognosis of patients with chronic hepatitis C-associated cirrhosis complicated by T2DM.

## Data Availability

All data produced in the present study are available upon reasonable request to the authors

## Author Contributions

**Data curation:** Xiaowu Wang, Yunyun Ding, Yan Liu, Tuantuan Li, Yi lang Zhu.

**Formal analysis:** Dong Wu, Xiaowu Wang, Yi lang Zhu, Yong Gao, Xiaojuan Wang.

**Methodology:** Dong Wu, Xiaowu Wang

**Project administration:** Yong Gao, Xiaojuan Wang.

**Writing – original draft:** Dong Wu, Xiaowu Wang, Yong Gao, Xiaojuan Wang.

**Writing – review & editing:** Dong Wu, Xiaowu Wang, Yi lang Zhu, Yong Gao, Xiaojuan Wang.

